# Loss of Y chromosome in Alzheimer disease patients co-occurs with clonal hematopoiesis defined by post-zygotic point mutations outside canonical CHIP driver genes

**DOI:** 10.1101/2025.06.05.25329040

**Authors:** Edyta Rychlicka-Buniowska, Daniil Sarkisyan, Monika Horbacz, Bożena Bruhn-Olszewska, Kinga Drężek-Chyła, Mikołaj Koszyński, Hanna Davies, Ulana Juhas, Magdalena Wójcik-Zalewska, Alicja Klich-Rączka, Lena Kilander, Martin Ingelsson, Karolina Bukowska-Strakova, Kazimierz Węglarczyk, Maciej Siedlar, Jarosław Baran, Janusz Ryś, Arkadiusz Piotrowski, Natalia Filipowicz, Jan P. Dumanski

## Abstract

Loss of Y chromosome (LOY) and clonal hematopoiesis of indeterminate potential (CHIP) are common age-related events associated with multiple adverse outcomes in the elderly. While LOY has been associated with higher risk of Alzheimer disease (AD), CHIP has been suggested to perform a protective role against AD. Moreover, the co-occurrence of CHIP and LOY is debated. We performed deep whole-exome sequencing of FACS-isolated CD4^+^ T lymphocytes, NK and myeloid cells from men with AD and controls exhibiting either LOY or retention of Y chromosome (ROY). We found 39 sequence variants in known (canonical) myeloid driver genes of clonal hematopoiesis (MD-CH) and known lymphoid driver genes (LD-CH), and maximally 14 (35%) of these could co-exist with LOY within the same clone. We further describe 192 unknown drivers of clonal hematopoiesis (UD-CH), which were markedly enriched in AD-LOY individuals (odds ratio=4.8, Benjamini-Hochberg-adjusted p=0.041), and over 20% of these variants were protein-truncating. In myeloid cells, the total burden of all detected drivers correlated with the percentage of LOY cells (Spearman ρ=0.52, adjusted p=0.00041). In conclusion, our findings suggest that LOY acts as the primary driver of clonal hematopoiesis in AD by seeding myeloid clones. These clones may subsequently accumulate additional, often truncating, UD variants, while most canonical CHIP mutations arise independently of LOY. Our study delineates distinct yet partially overlapping clonal architectures for LOY and CHIP in late-onset AD and underscores LOY-driven myeloid expansion as a potential contributor to disease pathogenesis.

## Introduction

Aging is accompanied by an accumulation of post-zygotic (somatic) mutations, particularly in hematopoietic stem cells (HSCs), which are estimated to acquire up to 0.858 exonic mutations per year, summing up to about 1.4 million protein-coding mutations in a 70-year-old individual^1,2^. When an HSC harboring such alterations gains a selective advantage, it gives rise to an expanded population of cells, in the process known as clonal hematopoiesis (CH). Clonal hematopoiesis of indeterminate potential (CHIP) specifically refers to CH with mutations accumulating in myeloid malignancy-associated genes^3–5^. These CH/CHIP mutations are typically present in blood cells with a variant allele frequency (VAF) of at least 2% in the driver genes. However, variants below this threshold can also have adverse outcomes^6^. Both CH and CHIP refer to individuals, who do not meet diagnostic criteria for hematologic malignancies. CHIP is not only associated with higher risk of acute myeloid leukemia^7,8^, but also with cardiovascular disease, all-cause mortality^9,10^, and chronic kidney disease^11^.

Loss of Y chromosome (LOY) is the most common post-zygotic aberration, occurring in normal hematopoietic cells^12,13^ and exhibiting temporal dynamics^14,15^. Hematopoietic LOY is detected in ∼20% of men aged 40–70^16^ and increases with age - from 5% in men younger than 50 years to 40% in those older than 65 years, reaching ∼60% by 93 years^17^. LOY has also been found in other tissues, albeit at substantially lower frequencies^18–20^. Initial reports correlated LOY to all-cause mortality, elevated risk of non-hematologic cancers and Alzheimer disease (AD)^21,22^. Subsequent studies have linked it to a broader spectrum of disorders (summarized in Table 1 of Bruhn-Olszewska et al.^23^).

**Table 1.**
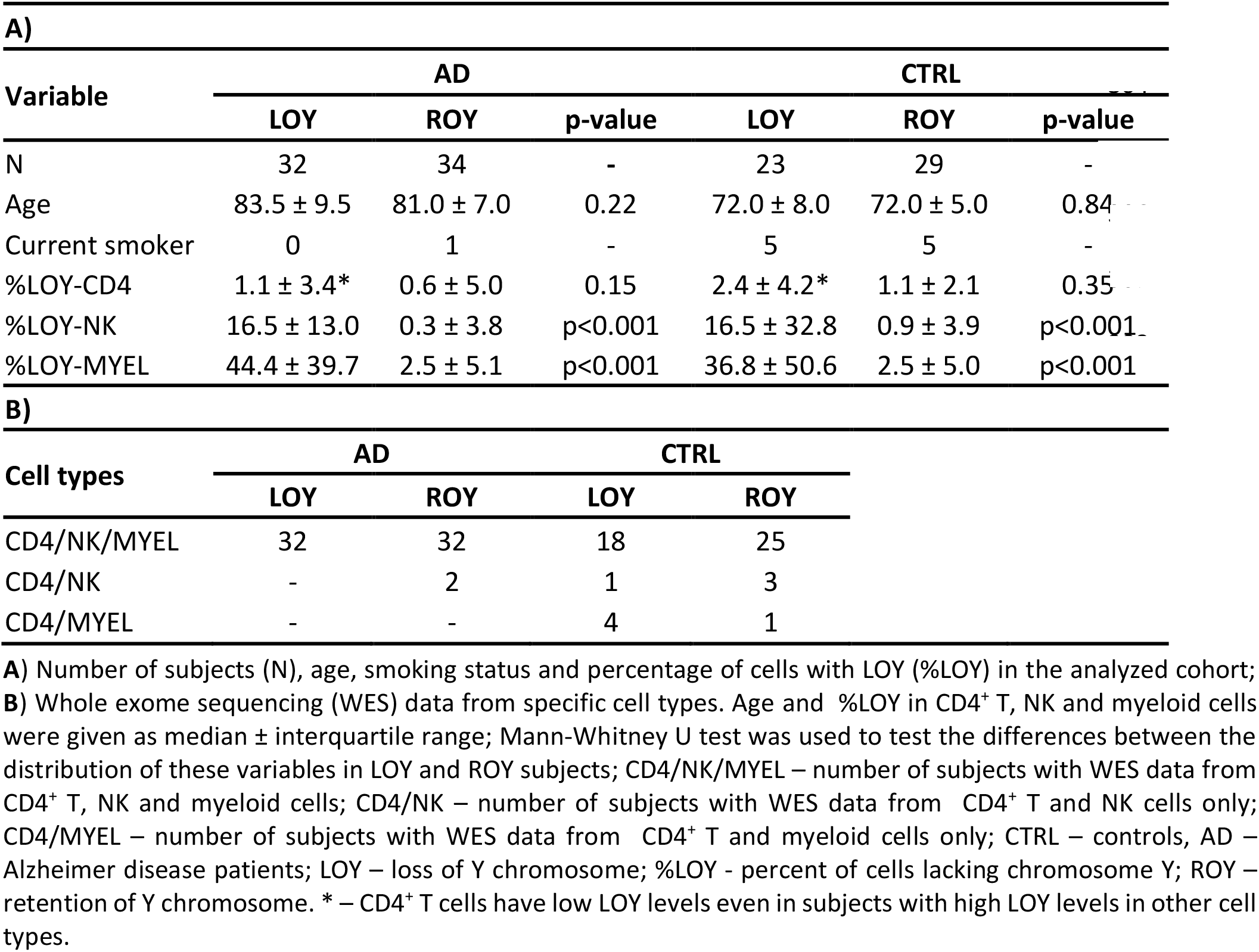
Basic cohort characteristics.

Since CHIP and LOY are common and age-related clonal events, investigating their co-occurrence and potential interaction emerged as a natural and important direction to understand age-associated clonal dynamics. LOY in bone marrow CD34+ cells was found in subjects with myelodysplastic syndrome^24,25^. In monocytes, LOY frequently co-occurs with pathogenic CHIP variants in individuals free of hematologic disorders^26^. Furthermore, age-related

CHIP in *TET2, TP53* and *CBL* genes has been associated with LOY occurring at clonal fractions exceeding 30%^27^, while large-scale population studies indicate that CHIP and LOY share germline genetic risk alleles^28^. Conversely, others describe reduced LOY in CHIP-positive individuals^29^ or even no co-occurrence between LOY and CHIP^30,31^.

Both CHIP and LOY are linked to immune dysregulation^12,19,32–35^, raising interest in their roles in neurodegenerative diseases. CHIP may confer neuroprotective effects by modulating systemic inflammation, potentially through altered microglial activity or other immune-related pathways slowing the progression of AD^36^. Conversely, CHIP has been linked to increased risk of vascular neurodegenerative conditions and amyotrophic lateral sclerosis^37^ with common driver mutations in *DNMT3A* and *TET2* concomitant with higher odds of developing Parkinson’s disease^38^ and multiple system atrophy^39^. LOY has likewise been implicated in AD pathogenesis, with evidence of its presence in leukocytes and brain cells^22,40,41^. The highest frequency of LOY-associated transcriptional changes in immune-related genes occurs in NK cells^22^, whereas LOY-related alterations in DNA methylation have been reported in granulocytes and monocytes^42^. Notably, LOY has also been detected in microglia from AD patients’ brains, suggesting its role in male-specific neurodegeneration^43^.

Given the accumulation of CHIP and LOY and their link to neurodegeneration, we set out to investigate their co-occurrence and potential pathogenic contribution to AD. We applied fluorescence-activated cell sorting (FACS) to isolate subpopulations of lymphoid (CD4^+^ and NK) and myeloid (granulocytes or monocytes) cells from AD patients and controls, stratifying individuals based on LOY or retention of Y chromosome (ROY). To analyze CH beyond the canonical CHIP driver genes, we performed high-coverage whole-exome sequencing (WES) on sorted fractions, interrogating 432 well-characterized myeloid and lymphoid driver genes alongside other protein-coding genes.

## Methods

### Study participants

Sixty-six AD patients were recruited at the Adult Psychiatry Clinic of the University Clinical Center in Gdańsk, the Clinic of Internal Diseases and Gerontology of the Jagiellonian University in Kraków and the Memory Clinic at Uppsala University Hospital. Fifty-two controls (CTRL), recruited from the general population of Gdańsk, Kraków and the EpiHealth study in Uppsala, met inclusion criteria of being over 65 years old with no self-reported history of cancer or dementia. Subjects were recruited between 2015 and 2022. Written informed consent was obtained from all participants. The study was performed in accordance with the Declaration of Helsinki and approved by the relevant local bioethical committees.

### Isolation of target leukocyte subpopulations and determination of LOY

CH was assessed in CD4+ T and NK cells (lymphoid lineage) and in granulocytes or monocytes (myeloid lineage). These cell subpopulations were isolated from peripheral blood with FACS, according to published protocols^12,34^. The proportion of cells with LOY (%LOY) in each cell type was determined at the DNA level using mLRR-Y values from Illumina SNP arrays^21^ and/or droplet digital PCR (ddPCR) to study the *AMELX*/*AMELY* genes^14^. Cell subpopulations with ≥15% of cells lacking the Y chromosome were classified as LOY, and subjects were assigned to the LOY group if any target subpopulation met this criterion.

### Whole exome sequencing (WES) and post-zygotic variants identification

WES was performed using SureSelect Human All Exon V7 kit (Agilent) with a low-input protocol to achieve an average on-target coverage of 150X. Post-zygotic single nucleotide variants (SNVs) and small indels were identified according to GATK4 best practices, and variant calling was performed separately using Platypus v0.8.1.1. A list of 97 myeloid and 335 lymphoid driver genes was curated based on current CH research (Tables S1 and S2)^5,26,27,29,44– 46^ with all other protein-coding genes categorized as unknown drivers.

### Statistical analysis

Logistic regression adjusted for age and age^2^ was used to assess the association between LOY and CH types in AD and CTRL donors, with the association strength expressed as OR with 95% CI. Differences in the distribution of age, %LOY and VAFs were evaluated using the Mann-Whitney U test. Spearman’s rank-based correlation was calculated within each cell type between the total CH burden (sum of variants’ VAFs) and the LOY level, and visualized with a least-squares line fitted to the ranked values of these two variables. All p-values were adjusted for multiple comparisons using Benjamini-Hochberg (BH) method.

### Gene set enrichment analysis (GSEA) and mRNA expression analysis

GSEA was conducted using R package clusterProfiler v4.14.6 with default settings, except that the minimum gene set size was set to 5. Genes with post-zygotic SNVs were ranked by classification, frequency and predicted functional impact. The analysis integrated multiple annotation databases, including Gene Ontology Biological Process (GO_BP) and Molecular Signatures Database (MSigDb 2024.1.hs). Gene sets with permutation-based BH adjusted p-values <0.05 were considered significantly enriched. Additional methods are provided in the *Online Supplementary Appendix*.

## Results

### The landscape of CH variants versus LOY status

Our aim was to analyze CH in peripheral leukocytes from lymphoid and myeloid lines collected from AD and CTRL individuals, stratified by LOY or ROY. Cell subpopulations with ≥15% of cells lacking the Y chromosome were classified as LOY. CH was defined by post-zygotic single nucleotide variants (SNVs) and small indels, detected using WES with deep coverage (median on-target coverage 136, range 62-200; Table S3). Variants were classified as post-zygotic if detected in one or more, but not all, cell types from the same individual. Based on the affected genes, variants were categorized as myeloid drivers (MD-CH), lymphoid drivers (LD-CH), or unknown drivers (UD-CH). MD-CH and LD-CH variants affected genes associated with myeloid and lymphoid malignancies^5,26,27,29,44–46^, respectively, while UD-CH comprised variants in all other protein-coding genes. The lists of the genes queried in this study are presented in the Supplementary Tables S1 and S2, respectively. .

The analyzed cohort included 66 AD patients (32 LOY, 34 ROY) and 52 CTRLs (23 LOY, 29 ROY) (Tables 1A and S4). Each subject contributed at least two cell types from lymphoid and myeloid lineages; 107 (90,7%) had all three fractions studied (Table 1B). In 193 samples LOY level was estimated using mLRR-Y, for 133 samples we used ddPCR assay for *AMELY*/*AMELX* genes and 22 samples were analyzed using both methods, with ddPCR results preferred for LOY scoring (Table S4). Notably, the LOY levels were typically higher in myeloid and NK cells, while CD4^+^ T cells showed lower levels even among LOY-classified subjects, which is consistent with previous findings^12^.

The distribution of detected variants across the 118 analyzed subjects, stratified by disease status (AD vs. CTRL) and Y chromosome status (LOY vs. ROY), is shown in Figure 1 and Table S5. In total, 13 MD-CH variants (including one *SRSF2* variant occurring twice) were identified in 12 subjects (12/118; 10.2%) – four AD patients (4/66; 6.1%) and eight CTRLs (8/52; 15.4%). Most MD-CH variants were protein-truncating (9/13; 69%) and had high oncogenic potential based on COSMIC and/or ClinGen-CGC-VICC^47^ annotations (12/13; 92%). The recurrently affected myeloid driver genes were *TET2, DNMT3A* and *SRSF2* (Table 2A). No significant enrichment of MD-CH variants was observed in LOY subjects. LD-CH variants were detected in 11 subjects (11/118; 9.3%), including 5 AD patients (5/66; 7.6%) and 6 CTRLs (6/52; 11.5%), each carrying a unique variant, five of which (5/11; 45%) were protein-truncating. *KMT2D* was the only recurrently affected lymphoid gene and variants were exclusively observed in LOY subjects (Table 2A). Furthermore, 192 UD-CHs were identified in 48 subjects (48/118; 40.7%), including 25 AD patients (25/66; 37.9%) and 23 CTRLs (23/52; 44.2%). Among these, 40 (40/192; 20.8%) variants were protein-truncating, detected in 26 individuals. There were no UD genes recurrently affected across subjects. UD-CHss were enriched in AD-LOY (18/32; 56.2%) in comparison to AD-ROY patients (7/34; 20.6%). Overall, clonal hematopoiesis, defined by post-zygotic variants in any driver category (MD-CH, LD-CH, or UD-CH; collectively “Any-CH”), was detected in 52 individuals (52/118; 44%), including 26 AD (26/66; 39.4%) and 26 CTRL patients (26/52; 50%).

**Table 2.** Recurrently affected genes and subjects with cell-type-specific CH variants.

| **A)** |  |  |  |
| --- | --- | --- | --- |
| **CH type** | **Gene** | **LOY** | **ROY** |
| MD-CH | TET2 | 2 | 3 |
| MD-CH | DNMT3A | 1 | 2 |
| MD-CH | SRSF2 | 0 | 2 |
| LD-CH | KMT2D | 2 | 0 |
| **B)** |  |  |  |
| **Cell type** |  | **LOY** | **ROY** |
| CD4 |  | 2 | 4 |
| NK |  | 11 | 10 |
| MYEL |  | 7 | 4 |
| NK/MYEL |  | 19 | 6 |
| CD4/NK/MYEL |  | - | 2 |

**A)** Number of subjects with CH variants affecting known myeloid (MD-CH) and lymphoid (LD-CH) driver genes recurred in LOY and ROY groups; **B)** number of subjects with cell-type-specific CH variants: MD-CH, LD-CH and unknown driver genes (UD-CH).

**Figure 1.**
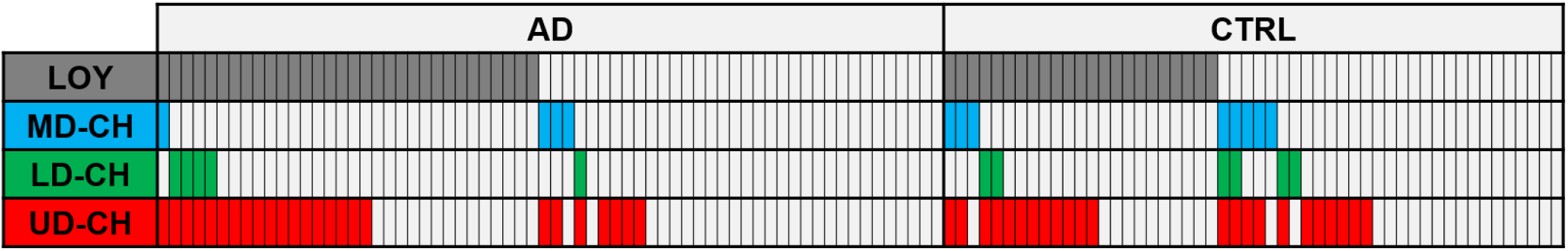
Landscape of clonal hematopoiesis (CH) in 118 analyzed subjects. Each type of CH variants is stratified by the disease status (AD patients vs. controls) and chromosome Y status (LOY vs. ROY). Each column represents one subject. Each gray rectangle in the first row represents the LOY subject. Each following row represents one type of CH variants, and the presence of variants in subject is indicated by a specific color: blue – known myeloid driver gene (MD-CH), green – known lymphoid driver gene (LD-CH), and red – unknown driver gene (UD-CH). AD: n=66 (32 LOY, 34 ROY); CTRL: n=52 (23 LOY, 29 ROY); AD – Alzheimer disease; LOY – loss of Y chromosome; ROY – retention of Y chromosome; CTRL – control.

### The association between LOY and CH in AD patients and controls

To assess if different CH variants were enriched in LOY subjects, we stratified AD and CTRL donors based on the chromosome Y status. The proportions of subjects with MD-CH, LD-CH, UD-CH and Any-CH in LOY and ROY groups were compared using logistic regression adjusted for age and age^2^. There was no significant association between LOY and MD-CH or LD-CH (Figure 2A and 2B). However, UD-CHs (OR=4.75, adjusted p=0.041) and Any-CH variants (OR=4.29, adjusted p=0.041) were significantly overrepresented in AD-LOY patients (Figure 2C and 2D).

**Figure 2.**
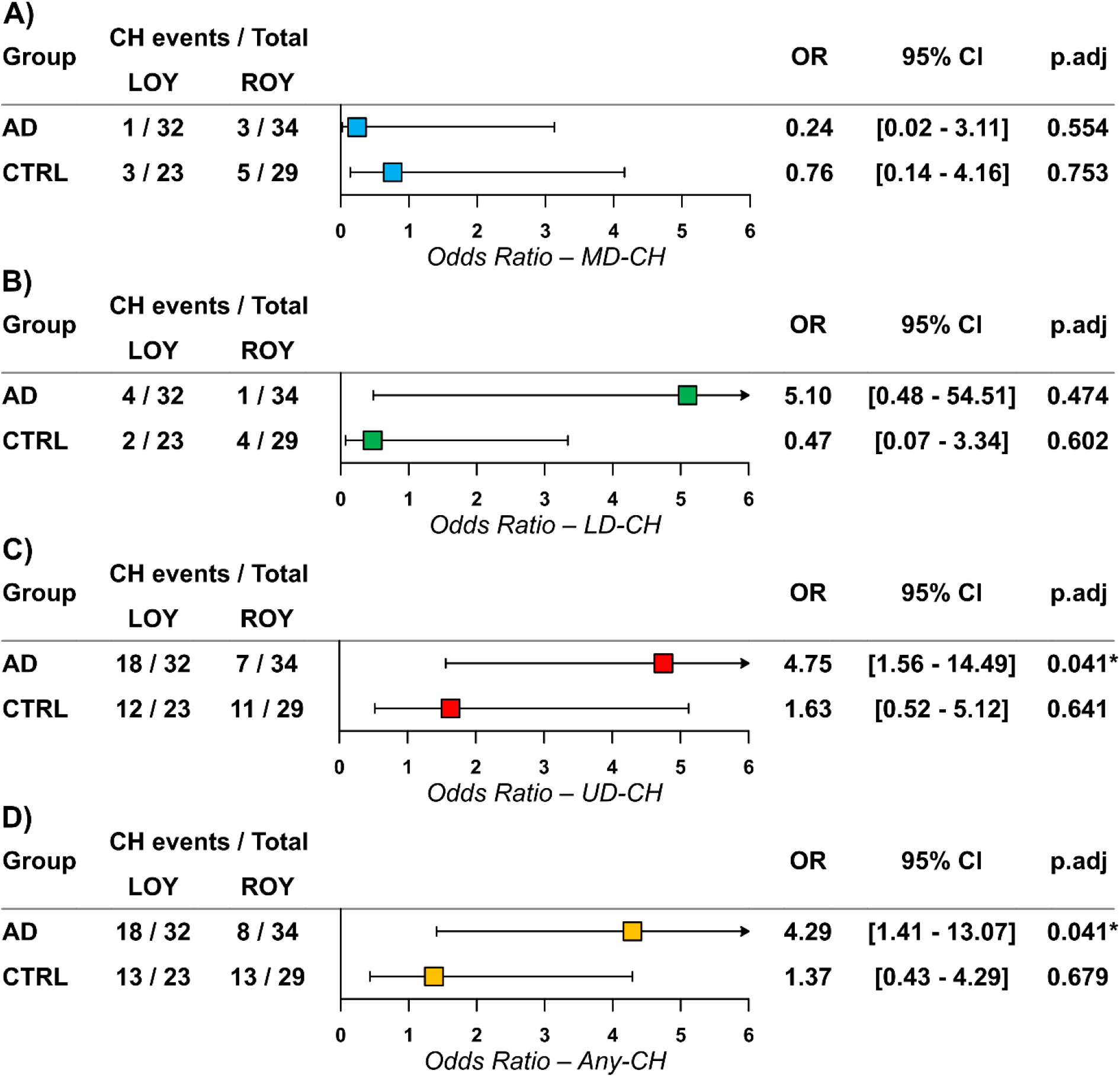
Association between LOY and CH types. A) LOY and MD-CH; B) LOY and LD-CH; C) LOY and UD-CH; D) LOY and all CH combined (Any-CH). Proportions of subjects with each CH type were compared between LOY and ROY groups within AD patients (AD) and controls (CTRL) using logistic regression adjusted for age and age^2^. Odds ratios (OR) with 95% confidence intervals (CI) are shown. P-values were Benjamini-Hochberg adjusted for eight comparisons (p.adj), with significant adjusted p<0.05 marked by an asterisk. Arrows indicate CIs extending beyond axis limits

To analyze the association between LOY and the VAFs of MD-CH, LD-CH and UD-CH variants, we stratified the cohort according to the chromosome Y status (Figure 3A). UD-CHs were analyzed separately in AD and CTRL group (Figure 3B). If several variants of the same CH type were detected in a subject, only the one with the highest VAF was included in the analysis. The VAFs of MD-CH variants were significantly lower in LOY subjects (adjusted p=0.020). The distributions of VAFs of LD-CH and UD-CH were similar in LOY and ROY individuals (Figure 3A). Although UD-CHss were enriched in AD-LOY group (Figure 2C), there were no significant differences in the distribution of their VAFs between AD-LOY and AD-ROY patients, similarly to the CTRLs (Figure 3B). The tendency of VAFs of MD-CH variants to be lower in the LOY group is also visible on Figure 4, where the percentages of cells (2xVAF) with different CH types are shown according to the %LOY in the analyzed cell fractions and the chromosome Y status on the subject level. While MD-CH variants in LOY subjects tend to cluster on the lower part of the x-axis, LD-CH and UD-CH variants are distributed across the entire scale, both in LOY and ROY group.

**Figure 3.**
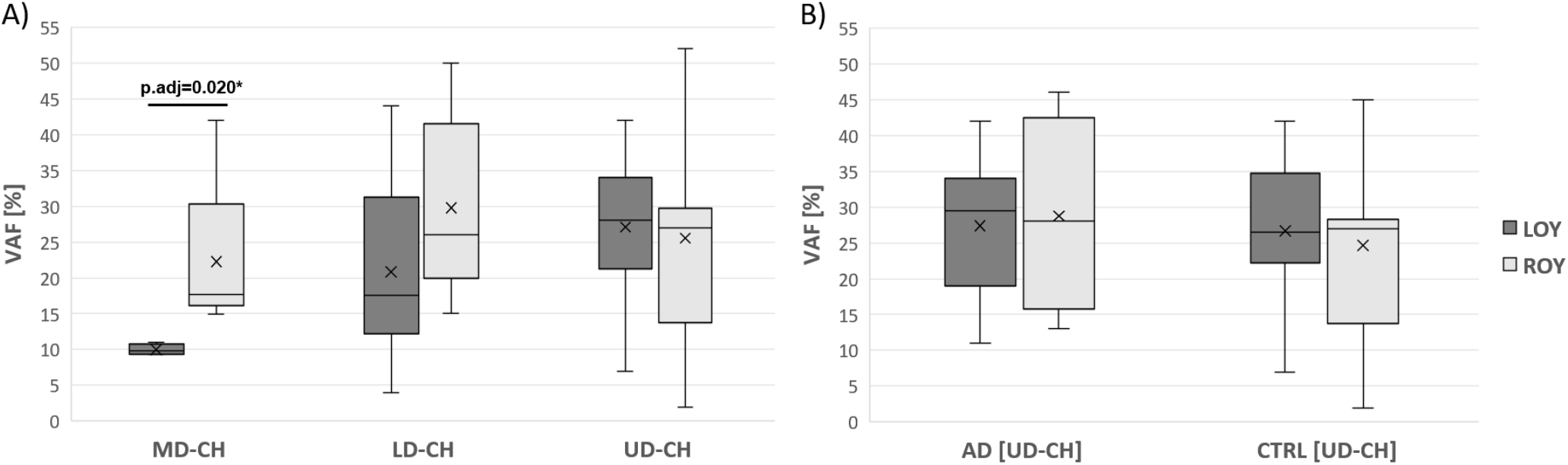
VAFs of different types of CH variants vs. LOY. The boxplots present the distribution of VAFs of variants detected in LOY and ROY subjects. A) VAFs of MD-CH, LD-CH and UD-CH variants in the cohort stratified according to the subject’s chromosome Y status only (AD patients and controls combined); B) VAFs of UD-CH variants in AD patients and controls. In case if more than one variant of the same CH type was detected in a subject, only the variant with the highest VAF was included in the analysis. The differences between distributions of VAFs in LOY and ROY subjects were tested using the Mann-Whitney U test. The p-values were adjusted for multiple testing using Benjamini-Hochberg method assuming 5 tests. Statistically significant adjusted p-value was marked with an asterisk. The line inside the boxplot represents the median; the cross inside the box represents the mean; the upper border of the box represents Q3; the lower border of the box represents Q1; whiskers extend from minimum to maximum. VAF – Variant Allele Frequency; AD – Alzheimer disease; LOY – loss of Y chromosome; ROY – retention of Y chromosome; CH – clonal hematopoiesis; MD – myeloid driver; LD – lymphoid driver; UD – unknown driver; Q3 – third quartile; Q1 – first quartile.

**Figure 4.**
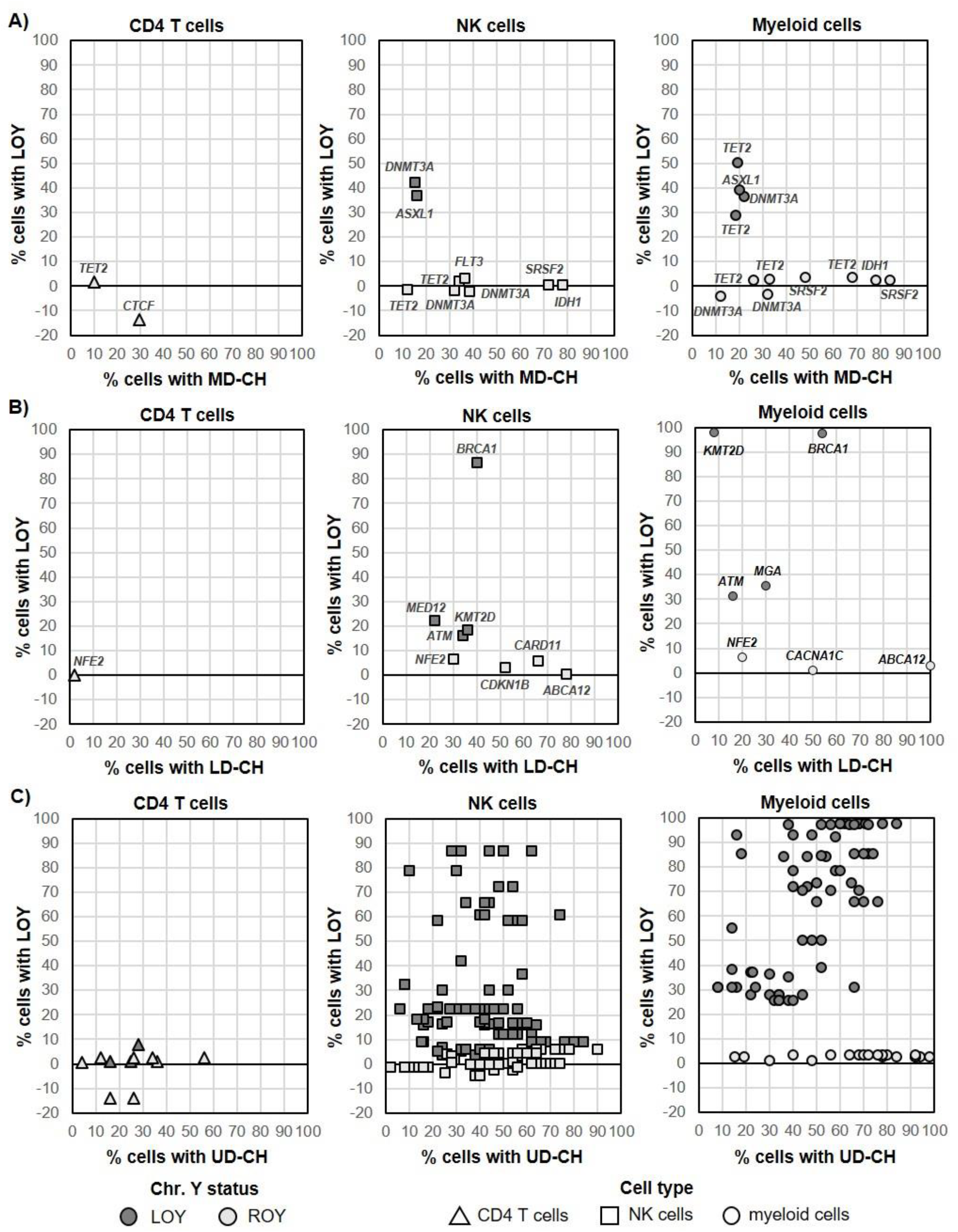
CH vs LOY across analyzed cell types. Scatterplots display the percentage of cells carrying each CH variant type (x-axis) vs. the corresponding percentage of cells with LOY (y-axis). (A) MD-CH vs. LOY in CD4^+^ T cells, NK cells and myeloid cells; (B) LD-CH vs. LOY in the same cell types; (C) UD-CH vs. LOY in the same cell types. Each point represents one variant detected in one subject. All variants detected in a given subject were shown. Gray background encodes Y chromosome status on the subject level (LOY ≥ 15% in any cell fraction); point shape denotes cell type. Gene symbols are annotated for MD-CH and LD-CH variants.

### Cell-type specific assessment of CH variants

To explore the cell-type specificity of CH variants, we assessed sharing of variants between sorted cell types (Table 2B). Two protein-truncating variants (MD-CH affecting *TET2* in ROY CTRL GK008 and LD-CH affecting NFE2 in ROY CTRL GK017), predicted to have high oncogenic potential based on COSMIC and/or ClinGen-CGC-VICC guidelines^47^, were detected in all three cell types. Both showed markedly lower VAFs in CD4^+^ T cells (0.05 and 0.01, respectively; Table S5) compared to NK and myeloid cells, where VAFs ranged from 0.10 to 0.17. Since this pattern suggested early clonal origin, likely at the level of hematopoietic stem or progenitor cells, with preferential expansion in NK and myeloid lineages, both variants were retained in the CH analysis as post-zygotic. Shared variants between NK and myeloid cells were observed in 25 subjects (19 LOY, 6 ROY), suggesting a common clonal origin with lineage-specific expansion. CH variants exclusive to NK cells were found in 21 subjects (11 LOY, 10 ROY). In contrast, CD4^+^ T cell–specific variants were rare, detected in only six individuals (4 ROY, 2 LOY).

The analysis of association between CH and LOY showed the total of 39 known MD/LD drivers, with 13 that possibly coexist with LOY: *ASXL1, DNMT3A, BRCA1, MED12, KMT2D* and *ATM* in NKs and *TET2, ASXL1, DNMT3A, KMT2D, BRCA1, ATM*, and *MGA* in myeloid cells (Figure 4A and 4B). Similarly, we found 65 and 67 UD-CH events in NK/myeloid cells, respectively, possibly co-occurring with LOY. The effect of LOY on total CH burden (a sum of VAFs of all post-zygotic variants detected in a sample) is presented in Figure 5. A significant Spearman correlation (ρ=0.52, adjusted p=0.00041) was observed in myeloid cells, but not in CD4+ T or NK cells, suggesting that LOY and CH tend to co-occur in granulocytes and monocytes, and LOY may be the primary driver of CH in myeloid lineage. On the contrary, CH in NK cells may be less frequently driven by LOY, as also supported by numerous variants with VAFs not proportional to %LOY (Figure 4, Table S5).

**Figure 5.**
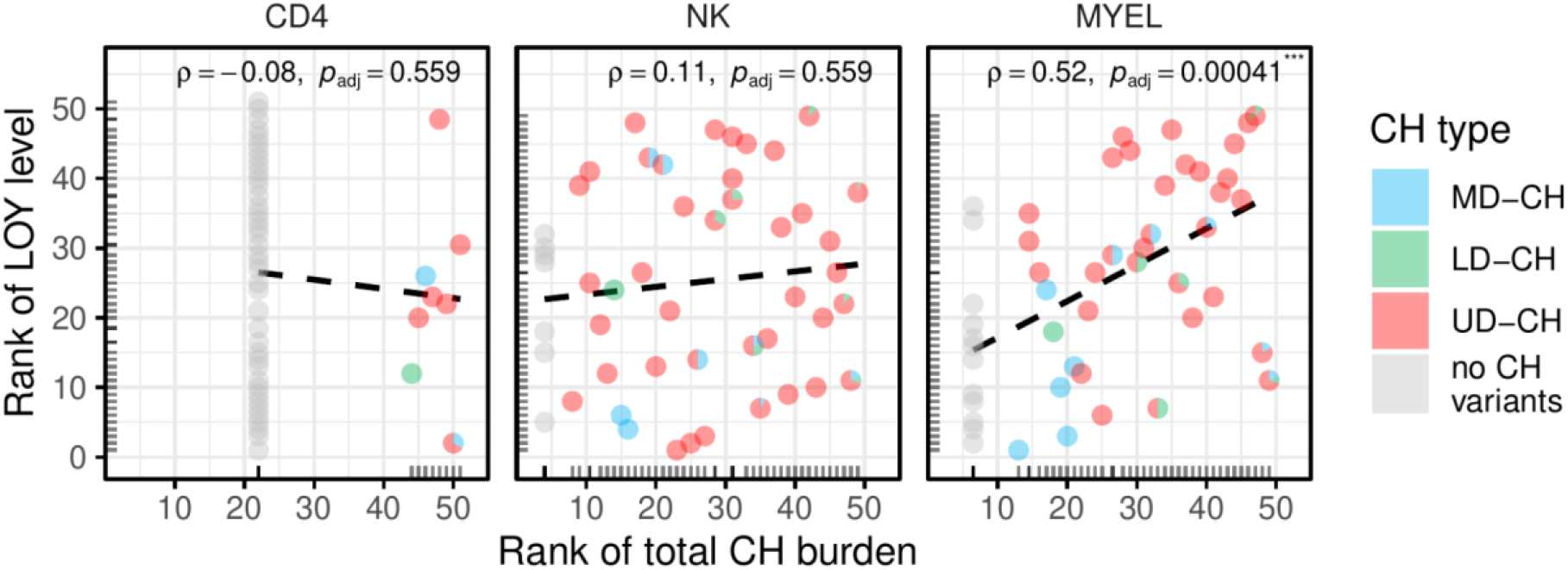
Total CH burden vs. LOY across cell types. For CD4^+^ T-cells, NK-cells and myeloid cells in respective facets, these scatterplots illustrate the strength and direction of the monotonic association between total CH burden (defined as sum of VAFs of all post-zygotic variants detected in a sample; x-axis) and LOY level (y-axis). Axes are equal-scaled, variables are converted to ranks within their respective cell-type, and bottom and left rugs indicate marginal rank distributions. Each subject is shown as a pie glyph depicting the proportional contribution of MD-CH (blue), LD-CH (green) and UD-CH (red) variants. Samples lacking detectable CH are rendered as solid grey pies. Dashed black lines are least-squares fits to the ranked data, visually representing Spearman’s correlation coefficient ρ and its BH adjusted p-values.

At the individual level, CH displays considerable complexity (Table S5). In the LOY group, clonal expansion appears to be driven by LOY in NK and myeloid cells in selected cases (e.g., M454), often accompanied by LD-CH or UD-CH. VAFs correspond to LOY levels in some individuals but not in others (e.g., GK037, M428), suggesting that LOY is not always the primary driver of CH. Other cases, like KAD054, further highlight the diversity in clonal architecture and variable contributions of LOY and post-zygotic variants across cell types. In ROY subjects, classical cases are observed where expansion is driven by oncogenic myeloid variants coexisting with variants of uncertain significance in lymphoid or unknown driver genes (e.g., subject M401). Some individuals (patient M394), exhibit NK-specific LD-CH variants with high oncogenic potential as a likely driver, alongside additional variants of other CH types. Donor GK008 showed a complex pattern with one specific MD-CH variant in myeloid cells and additional UD-CH variant(s) specific to CD4^+^ T or NK cells.

### Functional analysis of UD-CHs

Unknown driver variants(UD-CH) were found in 48 subjects and may contribute to clonal hematopoiesis (CH), especially in individuals without deleterious variants in known canonical drivers or LOY. As no recurrently mutated unknown driver genes were identified, we focused on their molecular functions and associated biological processes. To reduce the possibility of inclusion of variants of uncertain significance (VUS), we analyzed only protein-truncating and highly deleterious missense variants (M-CAP >0.025). Five main processes were identified: regulation of transcription by RNA polymerase II and DNA-templated transcription (12 genes: *HBP1, ABRA, CTBP2, ETV3, ZNF100, HNF1A, BMAL1, MTA2, NKAP, RBM10, MED12L, ADORA3*), regulation of splicing or transcriptional and post-transcriptional regulation of RNA (three genes: *DCDC2, PLEKHH1, CFAP96*), Rho GTPases and small GTPase-mediated signal transduction connected to cytoskeleton remodeling (six genes: *SYDE1, DMPK, MYH3, ABRA, RRAD* and *HEG1*), calcium homeostasis (six genes: *HEG1, RRAD, RYR1, GRIN2B, HTR5A, ATP2B3*) and ubiquitin-protein transferase pathways (two genes: *RBBP6* and *PDZRN3*).

To identify biological processes potentially disrupted by detected post-zygotic variants in all driver genes, we performed Gene Set Enrichment Analysis (GSEA). The analyzed cohort was stratified according to the chromosome Y status and disease status, which resulted in four groups: CTRL-ROY, CTRL-LOY, AD-ROY and AD-LOY. Results from two gene ontology databases are shown Table S6. In the AD-LOY group, significant enrichment (q<0.05) was observed for gene sets related to “immune system process” (NES=2.62, q=10-4) and “hemopoiesis” (NES=2.49, q<10-3), driven by genes including ASXL1 and ATM, suggesting perturbation of hematopoietic regulation and immune response. The AD-ROY group showed enrichment in regulatory gene sets, specifically targets of the miR-29 family (NES=2.21, q<0.01) and MYC transcription factor (NES=2.07, q=0.039). This enrichment was associated with mutations in epigenetic modifiers *DNMT3A* and *TET2*, indicating dysregulation of epigenetic and transcriptional networks. In the CTRL-ROY group, enrichment was significant for “leukemia” and “acute leukemia” terms (NES=2.12, q=0.03), driven by key myeloid malignancy-associated genes *DNMT3A, IDH1, SRSF2, and TET2*. This points towards a disruption of hematopoietic stem cell function predisposing to myeloid neoplasia. We did not detect any significantly enriched gene sets in the CTRL-LOY group. The analysis of mRNA expression of the 192 UD-CH genes in GeneCards database showed that the vast majority of them were clearly and highly expressed in immune cells. *SYTL5, TRIM71, IL1RAPL1, MAGEC1, SALL3, OR5K1, PCDHA12* expressed at intermediate-low level, while *FAM237A, OR5T1, AMER3* were at low levels, and *PRAMEF17* had no expression detectable above background in hematopoietic cells.

## Discussion

CH and LOY are common events in elderly, however their co-occurrence is a matter of debate. We employed FACS to isolate multiple hematopoietic cell lineages from each participant combined with deep WES. This approach enables robust detection of post-zygotic variants restricted to specific cell types and provides statistically meaningful LOY–point mutation co-occurrence estimates even in a modest-sized cohort, demonstrating the feasibility of scaling up this strategy. Querying public repositories with Ensembl Variant Effect Predictor^48^, we found that 57% of post-zygotic variants in our combined WES dataset are novel, 11% match catalogued post-zygotic only entries, and 32% bear mixed germline/post-zygotic annotations, underscoring our multi-lineage design as strongest point of the study. This support extending this framework to additional cell types and considerably larger cohort to refine variant catalogues and elucidate lineage-specific architecture.

We show that LOY infrequently co-occurs with variants in established drivers of myeloid and lymphoid malignancies, both in AD and CTRL cohorts (Figure 2A and 2B), suggesting that LOY alone does not drive the accumulation of highly pathogenic changes. We found altogether 39 sequence variants for known MD/LD drivers and maximally 14 (35%) of them could co-exist with LOY in the same hematopoietic clone (Figure 4). Additionally, the VAFs of detected MD-CH variants were significantly lower in LOY subjects (Figures 3A and 4), further supporting the lack of co-occurrence^30,31^ or even mutual exclusivity of LOY and CHIP^29^. The overall frequency of MD-CH in our cohort was 10.2%, irrespective of AD or LOY status, and recurrently affected MD genes were *TET2, DNMT3A* and *SRSF2*, which is consistent with the literature^7–9,27,38,44^. However, when stratifying the cohort according to disease status, we found that only 6.1% of AD patients had variants in myeloid drivers in comparison to 15.4% of CTRLs. Although this difference did not reach statistical significance, most probably due to the small cohort size, it is consistent with other studies, which report that CHIP is associated with reduced risk of AD^36^. It should also be stressed that even though AD patients in our cohort are ∼10 years older than controls, we do not see the accumulation of MD-CH with age in this group, which is in contrast to reports showing higher frequency of CHIP in older individuals^30,44^, and supporting the hypothesis of CHIP being the factor reducing the AD risk.

Similar to MD-CH, there was also no enrichment of LD-CH in LOY individuals, however, the OR for AD cohort was higher than for UD-CH, though not statistically significant, likely due to the rarity of these events and small cohort size (Figure 2B). The distributions of VAFs of LD-CH variants were similar in LOY and ROY groups (Figure 3A). Variants in genes driving lymphoid malignancies are usually classified as variants of uncertain significance (VUSs) with lower pathogenicity scores, and similar distribution of VAFs in LOY and ROY subjects could further support the hypothesis that LOY is not associated with highly pathogenic changes, but rather is followed by accumulation of VUSs. In contrast to MD-CH and LD-CH, UD-CH and Any-CH variants were significantly overrepresented in AD patients with LOY (Figure 2C and 2D), although there were no differences in the distributions of their VAFs between LOY and ROY individuals, both in AD patients and CTRLs (Figure 3A and 3B). Additionally, comparison of the variants identified in our study revealed overlap with previously reported LOY-associated transcriptional effects (LATE) genes: 18 UD-CH and 2 LD-CH genes had been reported to be dysregulated in myeloid cells of LOY subjects with AD^12,42^, while 51 UD-CH, 4 LD-CH, and 2 MD-CH genes corresponded to hypomethylated genes found in granulocytes and monocytes of AD-LOY patients^42^. These might suggest a role for LOY as a primary driver of these non-pathogenic passenger mutations in the AD cohort. It must be underscored that LOY at cellular fractions ≥30% has been associated with MD-CH, LD-CH and UD-CH in the general population^27^. However, determining whether LOY and CHIP truly co-occur within the same cell requires single-cell level analysis. In our cohort, LOY was detected in 15–97% of cells across 83 samples from 54 individuals (median for NK – 16.4%, and for myeloid cells – 38.2%). Given that the median VAF of mutations detected in NKs was 22%, and in myeloid cells 23% in LOY subjects, it is not possible to reliably infer the co-occurrence of LOY and CHIP within the same cell.

We also analyzed the UD-CHs for their pathogenicity and potential to drive CH. Overall, we uncovered 192 UD genes in 48 subjects, and these variants were predominantly observed in myeloid and NK cells. Twenty percent of UDs represent protein truncating mutations and these likely perturb immune homeostasis and hematopoietic regulation in which these proteins participate. The analysis of gene expression also showed that the vast majority of UD-CH genes are expressed in hematopoietic cells, supporting their potential relevance for CH. Further analysis of biological processes and molecular functions showed that in the AD-LOY cohort, post-zygotic mutations are significantly enriched in gene sets related to hematopoiesis, suggesting that LOY in combination with CH may perturb immune homeostasis. Moreover, shared UD-CHs were frequent between NK and myeloid cells especially in LOY subjects. Thus, LOY can act as a clonal driver in myeloid cells (and partially also in NKs), but its role is not universal and clonal expansions frequently occur independently of LOY. It is important to consider that some of the observed UD-CHs may represent passengers, expanding not through their own selective advantage but by clonal expansions initiated by LOY or other drivers. In this context, LOY may play a more central initiating role, potentially leading to the accumulation of additional mutations.

Consistent with the above, we show that there is a relationship between the total burden of observed CH variants and LOY in myeloid lineage, which is predominantly driven by UD-CHs (Figure 5). This supports the notion that CH is not driven by a restricted set of genes, but may involve a considerably broader range of mutations affecting regulatory processes in hematopoietic cells and this issue should be studied using sorted subpopulations of blood cells rather than using bulk DNA derived from all leukocytes. This could help in refining CH beyond already known driver genes. The lack of recurrence in uncovered affected genes between studied subjects, especially UD-CHs, further suggests heterogeneity in the mutational landscape of CH and points to the possibility of rare mutations contributing to CHIP on an individual basis. It is also important to acknowledge that some of the variants observed may represent passenger mutations, hitchhiking on clonal expansions driven by other, potentially unidentified, driver mutations. Distinguishing true drivers from passengers remains a major challenge and requires larger cohorts and functional validation. We should consider in this context that our cohort is relatively small and the issue of recurrence may be better resolved when the larger number of subjects is included. Overall, our approach to analysis of clonal expansion using deep WES in sorted cells suggests a complex picture of CH with potentially many more additional drivers, pointing to a still largely unexplored heterogeneity of CH-related sequence variants that may or may not be coexisting with LOY.

## Data Availability

All data produced in the present study are available upon reasonable request to the authors.

## Acknowledgements

We would like to thank all the patients and healthy controls for sample contribution and information provided in the questionnaires. We are grateful to Dr. Magdalena Koczkowska and Dr. Jakub Mieczkowski for the scientific support and advice.

## Notes

### Competing Interest Statement

JPD is a cofounder and shareholder in Cray Innovation AB. Other authors declare no competing interests.

### Funding Statement

This study was supported by grants from the Swedish Research Council, Cancerfonden, Hjärnfonden and Alzheimerfonden in Sweden to JPD and the Foundation for Polish Science under the International Research Agendas Program to JPD and AP (MAB/2018/6).

### Author Declarations

Independent Bioethics Committee for Research at the Medical University of Gdańsk, the Bioethical Committee of the Regional Medical Chamber in Kraków and the Uppsala Regional Ethical Review Board gave ethical approval for this work.

